# Advanced variant classification framework reduces the false positive rate of predicted loss of function (pLoF) variants in population sequencing data

**DOI:** 10.1101/2023.03.08.23286955

**Authors:** Moriel Singer-Berk, Sanna Gudmundsson, Samantha Baxter, Eleanor G. Seaby, Eleina England, Jordan C. Wood, Rachel G. Son, Nicholas A. Watts, Konrad J. Karczewski, Steven M. Harrison, Daniel G. MacArthur, Heidi L. Rehm, Anne O’Donnell-Luria

**Affiliations:** Program in Medical and Population Genetics, Broad Institute of MIT and Harvard, Cambridge, MA, USA; Center for Genomic Medicine & Analytic and Translational Genetics Unit, Massachusetts General Hospital, Boston, MA, USA; Division of Genetics and Genomics, Boston Children’s Hospital, Harvard Medical School, Boston, MA, USA; Science for Life Laboratory, Department of Gene Technology, KTH Royal Institute of Technology, Stockholm, Sweden; Genomic Informatics Group, University Hospital Southampton, Southampton, United Kingdom; Ambry Genetics, Aliso Viejo, CA, USA; Centre for Population Genomics, Garvan Institute of Medical Research and UNSW Sydney, Sydney, New South Wales, Australia; Centre for Population Genomics, Murdoch Children’s Research Institute, Melbourne, Australia

**Keywords:** predicted loss of function variants, pLoF, PVS1, gnomAD, variant classification, population data

## Abstract

Predicted loss of function (pLoF) variants are highly deleterious and play an important role in disease biology, but many of these variants may not actually result in loss-of-function. Here we present a framework that advances interpretation of pLoF variants in research and clinical settings by considering three categories of LoF evasion: (1) predicted rescue by secondary sequence properties, (2) uncertain biological relevance, and (3) potential technical artifacts. We also provide recommendations on adjustments to ACMG/AMP guidelines’s PVS1 criterion. Applying this framework to all high-confidence pLoF variants in 22 autosomal recessive disease-genes from the Genome Aggregation Database (gnomAD, v2.1.1) revealed predicted LoF evasion or potential artifacts in 27.3% (304/1,113) of variants. The major reasons were location in the last exon, in a homopolymer repeat, in low per-base expression (pext) score regions, or the presence of cryptic splice rescues. Variants predicted to be potential artifacts or to evade LoF were enriched for ClinVar benign variants. PVS1 was downgraded in 99.4% (162/163) of LoF evading variants assessed, with 17.2% (28/163) downgraded as a result of our framework, adding to previous guidelines. Variant pathogenicity was affected (mostly from likely pathogenic to VUS) in 20 (71.4%) of these 28 variants. This framework guides assessment of pLoF variants beyond standard annotation pipelines, and substantially reduces false positive rates, which is key to ensure accurate LoF variant prediction in both a research and clinical setting.

## INTRODUCTION

Loss of function (LoF) variants have important implications in human disease biology by either partial or complete loss of protein expression depending on the zygosity of the variant.^1,2^ Loss of protein expression is known to be caused by nonsense, frameshift, essential splice site, initiation codon variants, as well as structural variants spanning one or several exons, such as deletions and tandem partial gene duplications, with the first three (nonsense, frameshift, essential splice site) being annotated as pLoF (predicted LoF) by today’s standard annotation pipelines.^3,4^ However there are several mechanisms by which variants annotated as pLoF do not result in loss of protein expression, and careful interpretation beyond standard annotation is critical.^5,6^

Databases of human population genetic variation, such as the Genome Aggregation Database (gnomAD), enable us to refine our ability to interpret population genome sequencing data and assess variant pathogenicity.^7^ There are several possible reasons for evasion of true LoF that need to be considered, especially in disease genes. Previous studies have indicated that late truncating variants and variants that disrupt splicing at in-frame exons do not result in nonsense-mediated decay (NMD), but instead produce truncated protein products or in-frame deletions.^8,9^ Furthermore, all sequencing data is at risk for inclusion of sequencing artifacts, defined as variation introduced by a non-biological process such as read mis-mapping and base mis-calling.^10^

Current ACMG/AMP guidelines for sequence variant interpretation enable assessment of variants using criteria such as computational and predictive evidence, functional evidence, segregation, de novo evidence, population evidence, and allelic data.^11^ These guidelines are used worldwide to classify variants as pathogenic, likely pathogenic, uncertain, likely benign or benign. PVS1 is the ACMG/AMP evidence code for loss of function variation in a gene where LoF is a known mechanism of disease, and is the strongest weighted evidence in the ACMG/AMP curation process, which can result in pLoF variants being classified as pathogenic with only minimal additional evidence required. As such, it is clear that pLoF variants require a more in-depth assessment to accurately predict their effect. Therefore, the ClinGen Sequence Variant Interpretation (SVI) working group developed LoF interpretation guidelines, which outline how and when to count evidence for pLoF variants and apply strength modifications to the PVS1 code.^4^ These additional specifications go into detail regarding how to accurately identify falsely annotated pLoF variants that is not subject to NMD, such as LoF variants in the most 3’ exon of a gene that may produce a functional, albeit truncated, protein and therefore should not be given the full strength of PVS1.

Interpretation of pLoF variants is also important, and substantially more challenging, in the context of large population cohorts, where such variants have been consistently shown to be highly enriched for a wide variety of sequencing rescues, artifacts and annotation errors. The driver of this enrichment is Bayesian: there is a high prior probability that a pLoF variant observed in a known disease gene in a rare disease patient is real, but this probability is much lower for a similar variant observed in an individual ascertained at random from the population.^2,12^ As a result, studies applying careful curation to population cohorts have found consistently high rates of sequencing and classification errors.^7,13–15^ This enrichment is particularly striking in genes where true LoF variants are well-established to cause genetic disease that is not compatible with participation in common disease studies, thus not expected in population databases like gnomAD.^16–18^ Such high error rates complicate studies leveraging pLoF variants seen in large cohorts to explore human gene function, an approach that has proven extremely valuable for the identification and validation of potential therapeutic targets.^19^ While automated approaches to pLoF variant filtering remove a substantial fraction of errors,^7,18^ multiple studies have demonstrated the value of deep manual curation of pLoF variants to identify evasion modes and sequencing artifacts missed by these automated tools.^14,15,20^

Here we present an advanced LoF curation framework for interpreting pLoF variants (nonsense, frameshift, and essential splice site) that expands on current guidelines. This framework highlights when a pLoF variant may not be subject to NMD or when it is a potential technical artifact, the latter being especially useful when assessing data from population databases like gnomAD, where analytical validity cannot be verified. The framework considers three main categories: (1) predicted rescue by secondary sequence properties, (2) uncertain biological relevance, and (3) potential technical artifacts. We present the manual curation of all high quality pLoF (heterozygous) variants in 22 autosomal recessive (AR) disease genes in 141,456 individuals from gnomAD v2.1.1 using this framework. Additionally, we provide guidance on how to utilize this framework for applying PVS1 criteria and interpreting pathogenicity, in line with, and further building on, the ACMG/AMP and ClinGen SVI recommendations for PVS1 use.^4,11^

## MATERIALS AND METHODS

The framework was developed by defining established mechanisms of pLoF evasion and identifying potential technical artifacts as previously reported in the literature.^4,13^ A set of 22 AR disease genes with 1,113 pLoF variants was selected for manual curation based on pLoF annotation. The analysis included variants passing gnomAD quality control filters, excluding low confidence genotypes (depth <10, genotype quality <20, allele balance <20% for non-reference heterozygous variants) and excluding outliers of the random forest model that considers allele-specific annotations. Any variant annotated as pLoF by the Variant Effect Predictor (VEP; stop-gained/nonsense, essential splice acceptor/donor (-/+1-2), or frameshift variants) in either exomes or genomes for any transcript in gnomAD v2.1.1 (VEP version 85 using GENCODE v19 on GRCh37) was included for this analysis. Variants annotated as low-confidence by the Loss-of-Function Transcript Effect Estimator^7^ (LOFTEE; removing variants less likely to result in LoF) were excluded. Manual curation was then independently performed by two biocurators in a custom curation interface (https://github.com/macarthur-lab/variant-curation-portal), and any discrepancies were resolved by group discussion. Resources used for curation of pLoF variants included gnomAD variant and gene pages,^12^ UCSC genome browser,^21^ and SpliceAI for canonical splice variant interpretation.^22^ For transcript-level flags, variants were evaluated using “Basic Gene Annotation Set from GENCODE version 19” in the UCSC genome browser. A subset of pLoF variants curated as likely not LoF/not LoF were additionally assessed for effects on PVS1 using ACMG/AMP and ClinGen SVI guidelines.^4,11^ The correlation between variants that are predicted to evade LoF and benign variants in ClinVar was determined using variants that had at least one submission to ClinVar (479/1,113, https://ftp.ncbi.nlm.nih.gov/pub/clinvar/, January 5, 2022).

Each variant was assessed for evidence suggesting LoF evasion and potential technical artifacts by the rules that define the final verdict of LoF, likely LoF, uncertain LoF, likely not LoF, or not LoF (Table 1). For some rules the thresholds can be modified (conservative or lenient) depending on the overall aim of a curation project (Table 2). A conservative cut-off generates fewer false positive LoF/likely LoF but more false negatives (variants called as not LoF while in fact they cause true LoF), while lenient rules are more inclusive and will discard fewer variants as likely not LoF/not LoF but instead result in more false positive pLoF variants. Flags were applied conservatively to identify any variant potentially not causing LoF given that gnomAD, like any population database of genome and exome research data, is likely to be enriched for LoF artifacts.

**Table 1:** Rules that define LoF verdicts. When multiple rules apply to a pLoF variant the most impactful consequence is assigned. Specific thresholds and additional recommendations for a subset of rules, superscripts a-h, are found in Table 2.

| Error Type | LoF | Likely LoF | Uncertain | Likely not LoF | Not LoF |
| --- | --- | --- | --- | --- | --- |
| Predicted Rescue by Secondary Sequence Properties | Splice rescue that introduces stop codon | Translation re-initiation removing >25% coding sequence | Weak translation reinitiation<sup>e</sup> | Splice rescue with weaker predictions<sup>d</sup> | Splice rescue within stronger predictions<sup>d</sup> |
|  | Intron retention |  | In-frame and out of frame rescue events | Variant falls within overhang exon | MNV that results in a missense/synonymous variant |
|  |  |  |  | Strong translational re-initiation<sup>e</sup> | Frame-restoring indel |
|  |  |  |  |  | In-frame exon skipping (according to splicing prediction) |
| Uncertain Biological Relevance | Minority of transcripts with pext at maximum for gene<sup>g</sup> | Minority of transcripts with pext close to maximum for gene<sup>g</sup> | Minority of transcripts with pext <50 % of the maximum of the gene (pext > 20% of maximum)<sup>g</sup> | Weak exon conservation with pext <50% of the maximum of the gene (pext > 20% of maximum)<sup>g</sup> | Pext < 20% of maximum for the gene<sup>f</sup> |
|  | Weak exon conservation with pext at maximum for gene<sup>g</sup> | Weak exon conservation with pext close to maximum for gene<sup>g</sup> |  | Nonsense variant in overprinted transcript | Splice variant not supported by pext<sup>f</sup> |
|  |  |  |  |  | Variant terminates within the last exon or last 50bp of the penultimate exon<sup>h</sup> |
| Potential Technical Artifacts | No read data for splice and nonsense variants | Genotyping errors above threshold<sup>a</sup> | No read data for frameshift variants | Genotyping errors below threshold<sup>a</sup> |  |
|  | Strand bias | GC rich region |  | Homopolymer<sup>c</sup> |  |
|  |  | Low complexity sequence |  | Complex mapping errors<sup>b</sup> |  |
|  |  | Minor mapping errors<sup>b</sup> |  |  |  |

LoF, loss of function; MNV, multi-nucleotide variant; pext, proportion expressed across transcripts; bp, basepairs.

**Table 2:**
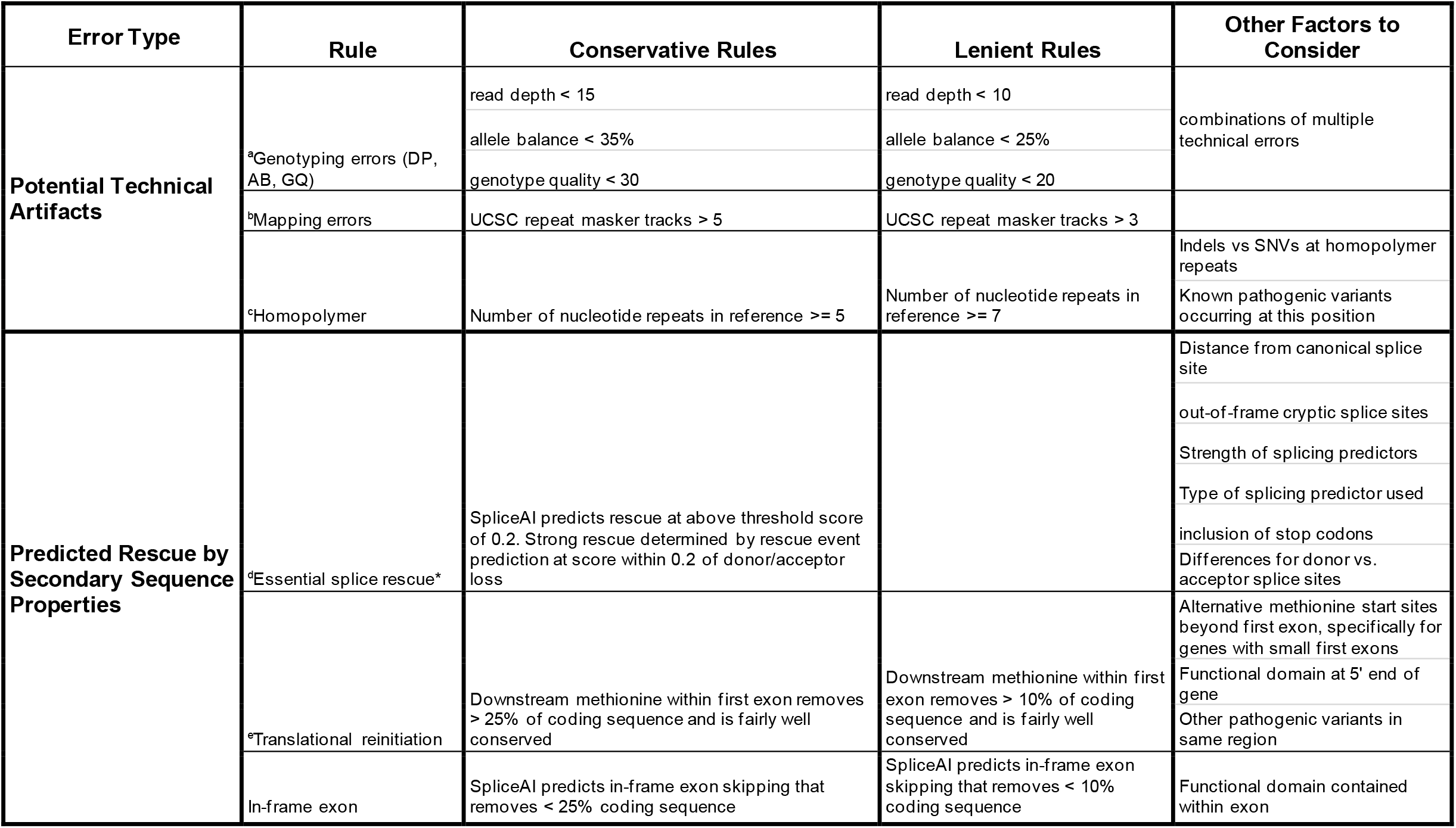
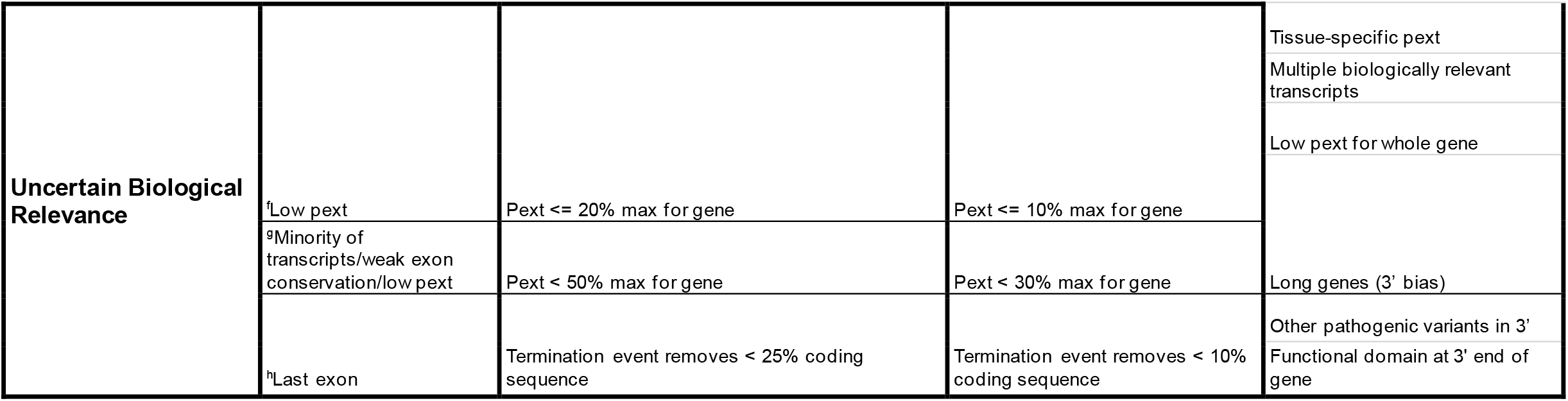
Specifics for the different LoF curation rules from table 1 applied depending on the type of curation (conservative vs. lenient). Each rule includes a list of other factors to consider when defining a framework for LoF curation. SNVs, single nucleotide variants; pext, proportion expressed across transcripts; bp, base pairs. *Essential splice rescue flag relies on the prediction of in silico tools, thus the recommended threshold for that tool applies. For SpliceAI that is 0.2, with no lenient threshold.

## RESULTS

### Evidence suggesting LoF evasion determines final verdict

Each pLoF variant is assessed for evidence of LoF evasion and assigned flags according to rules (Table 1) subdivided into three categories: (1) predicted rescue by secondary sequence properties, (2), uncertain biological relevance, and (3) potential technical artifacts (Figure 1). The combination of flags is used to determine pLoF verdict: (1) LoF (2) likely LoF, (3) uncertain LoF, (4) likely not LoF, or (5) not LoF. The visualization of read data to assess variant quality and potential rescues (such as frame-restoring indels) is essential to this protocol, thus, frameshift variants without read data are classified as “uncertain” unless additional evidence of the variant suggested LoF evasion or predicted the variant as a potential technical artifact.

**Figure 1:**
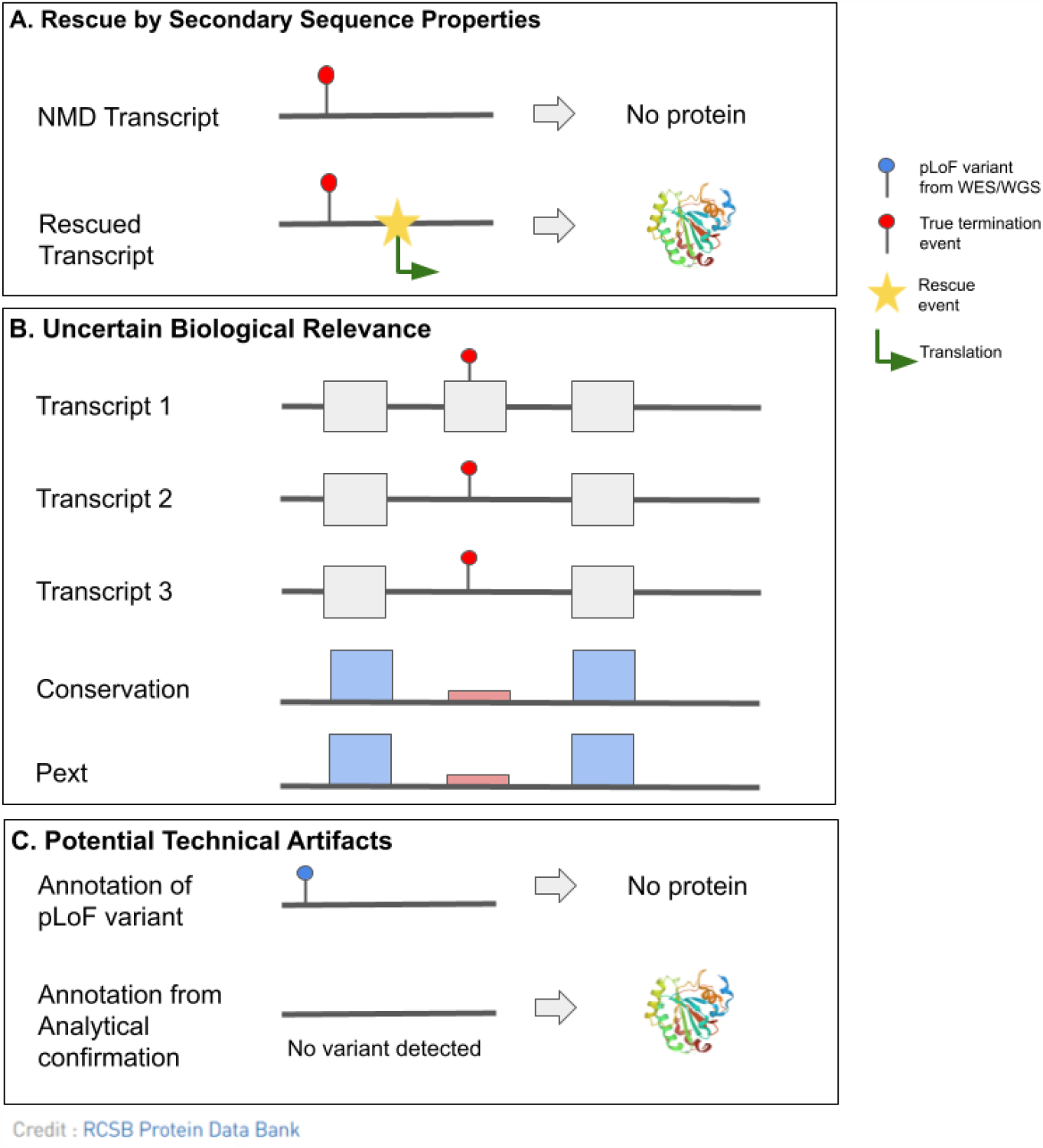
Schematic showing the main categories of flags in pLoF interpretation. (A) LoF evasion as a result of a predicted rescue by secondary sequence properties. A termination event near a source of rescue allows for translation of the sequence into protein and escapes NMD. (B) A termination event within an exon of uncertain biological relevance is predicted to evade loss of protein expression. Uncertain biological relevance can be identified here by a combination of the location of the termination event within a minority of transcripts, weakly conserved exon relative to surrounding region, and low mean pext score, suggesting that the affected exon is in fact of low biological importance. (C) Potential technical artifacts where analytical confirmation is needed to confirm the variant.

#### Predicted rescue by secondary sequence properties

Rescue flags are assigned to pLoF variants that are predicted to be rescued by a secondary sequence property such as an **in-phase multi-nucleotide variant (MNV**), **frame-restoring indel, essential splice site rescue, in-frame exon skipping, translational reinitiation**, and **overhanging exon**.^23–30^ Since standard variant annotation pipelines do not assess the variant in the context of the surrounding sequence, these variants will be called as pLoF despite nearby rescues.

**MNV**s refer to multiple SNVs found within the same codon and haplotype, that have arisen either as a single mutational event or as multiple coincidental mutations. MNVs often have a different effect on the protein sequence in aggregate than the same variants considered individually,^25,31,32^ but existing variant annotation pipelines consider all SNVs as independent events, resulting in errors in impact prediction of MNVs (Supplementary Figure S1). **Frame-restoring indels** also rescued by the presence of another variant on the same haplotype; for example one variant annotated as causing a frameshift can be rescued by one or several nearby indels, with the aggregate impact being an in-frame indel (potentially with some intervening sequence resulting in multiple missense substitutions) rather than a frameshift (Supplementary Figure S2).

Essential splice site variants cause LoF by disrupting splicing, typically resulting in usage of a cryptic splice site and/or exon skipping leading to introduction of an early termination codon.^33,34^ Using established splice predictors like SpliceAI can help predict the effect of variation at an essential site, *i*.*e* using an up or downstream cryptic splice site that is either in-frame or out-of-frame, exon skipping, intron retention, or a combination of these events. Out-of-frame cryptic splice sites will in most cases result in introduction of an early termination codon and NMD, while **in-frame cryptic sites** result in an in-frame indel without a loss of protein expression. Of note, for in-frame cryptic splice sites resulting in partial intron retention, the intronic sequence needs to be assessed for inclusion of termination codons that could result in early truncation and NMD. Essential splice site variants at the border of an **in-frame exon** can also result in an in-frame deletion of that exon rather than introducing an early termination codon and NMD, which can also be predicted by SpliceAI.

Other types of transcript rescue include **translational reinitiation** and **overhang exons**. The translational reinitiation flag is assigned to variants that have a nearby in-frame methionine downstream of the termination event that may re-initiate translation. The overhang exon flag is assigned to variants that fall in an exon extension (Supplementary Figure S3). Overhang exons are often weakly conserved, have a lower pext score, and fall in a minority of coding transcripts. Variants that fall within overhang exons are considered rescued by splicing out the overhanging sequence through essential splice sites of other transcripts and thus are predicted to evade LoF.

#### Uncertain Biological Relevance

Variants of uncertain biological relevance are expected to result in NMD within at least one transcript, but their effect on the overall protein expression is not predicted to have a biological impact. Specific flags include **minority of transcripts, weak exon conservation, low pext, overprinting**, and **last exon**. These flags highlight the requirement for in-depth interpretation of the variant, transcript, and conservation.^35–38^

Proportion expressed across transcripts (pext) scores, available on the gnomAD browser gene page, inform the relative per-base expression across transcripts in GTEx tissues.^18^ Variants that fall in **low pext** regions, defined as an exon with mean pext value <20% of the maximum pext across the gene, are often in biologically dispensable alternative transcripts (Supplementary Figure S4). It is of note, that most splice variants fall outside the coding region, and in those cases the pext score will always be 0. However, the biological relevance of the splice site can be interpreted by looking at the score of the adjacent exon; if the adjacent exon has a low pext score it is likely biologically dispensable. We also used the absence of a drop in the pext score at an annotated splice site in a transcript as evidence that it is likely a splice site (and transcript) of low biologic relevance (Supplementary Figure S5).

Variants that are annotated as pLoF in a **minority of coding transcripts** across the gene need to be assessed for whether or not they fall in the most biologically relevant transcript (e.g. MANE Select and MANE Plus Clinical are recommended for GRCh38). In genes with multiple transcripts, transcript expression in a specific tissue or the mean expression across tissues might be a useful indicator of biological relevance. In this protocol, we flagged variants occurring in <50% of coding transcripts. Likewise, a pLoF variant located in a **weakly conserved exon** may indicate that loss of that exon does not impact gene function. Some variants are located in exons with slightly lower pext scores (∼50% of gene maximum), a minority of transcripts and/or weakly conserved exons, implying that the exon itself is less relevant to gene function. The combination of these flags (minority of transcript, weak exon conservation, and low pext) can be used to inform the biological relevance of the transcripts in which a pLoF variant is annotated. Of note, lack of a pext score should not be used as evidence for or against LoF evasion if the variant is located in a transcript that was not included when pext scores were generated and thus, is not represented in gnomAD. It is also important to consider that GTEx gene-expression data is derived from adult postmortem tissues, which may not accurately represent gene-expression during early development^39^ and not all tissues are available (e.g. inner ear).

The **overprinting** flag is applicable for a transcript with an unconserved alternate open reading frame (ORF; Supplementary Figure S6). Overprinting has been described as a means for *de novo* gene birth and is widely reported in viral DNA, and more recently in plants and animals.^40–42^ A variant that is annotated as pLoF only in the “novel” overprinted transcript will likely have a different annotation (missense/synonymous) in the ancestral frame and is therefore not considered to cause LoF in the primary gene annotated at the locus.

Variants that terminate in the last exon of a gene or the last 50-55 bp of the penultimate coding exon typically result in truncated protein products due to NMD escape.^43,44^ Therefore pLoF variants assigned with the **last exon** flag are not predicted to result in lost protein expression but rather the expression of the truncated protein, which may or may not have a deleterious effect on protein function and needs further assessment. LOFTEE (v1.0.3) does not flag by NMD location and instead by GERP score of the affected region of the protein such that a number of pLOF variants predicted to escape NMD are not annotated as such by LOFTEE.^7^ Furthermore, not all genes are subject to NMD, and those that are not subject to NMD need to be interpreted differently. This is unknown for most genes and in the absence of other knowledge, our default assumption is to expect NMD.

#### Potential Technical artifacts

Technical flags are assigned to pLoF variants that are likely artifacts from sequencing data rather than true variants, and include **genotyping, mapping**, and **homopolymer flags**. Technical flags are used for variants in regions where the confidence of finding a real LoF variant is decreased by the region-quality and where there is a higher rate of false positive variant calls in exome and genome capture.^45–50^

**Genotyping flags** are assigned to variants with a skewed allele balance, low read depth, low genotype quality, or that fall in low complexity or GC rich regions, or demonstrate strand bias (variants called predominantly on the forward or reverse strand; Supplementary figure S7). **Mapping flags** are assigned to variants that fall in a region of the genome where there are known mapping difficulties due to repetitive genome wide sequences, and variation in these regions might be a result of mismapped reads (Supplementary Figure S8). UCSC genome browser’s repeats tracks can be used to identify regions that are likely to be mismapped. **Homopolymers**, a sequence of consecutive identical bases, are enriched for false positive indels due to polymerase slippage during PCR amplification. Slippage results in inaccurate reports of repeat length, which are then incorrectly annotated as frameshift variants.^51–55^

### Curation framework predicts 27% of pLoF variants do not result in LoF

Curation was performed on 1,113 pLoF variants in 22 AR disease genes determined high-confidence by LOFTEE. LOFTEE pre-filtered 143 variants as low-confidence mostly due to their location in the end-truncating region of the gene (Supplementary Figure S9).^7^ Of the 1,113 LOFTEE high-confidence pLoF variants, 304 (27.3%) were interpreted as likely not LoF/not LoF, 42 (3.8%) were interpreted as uncertain LoF, and 767 variants (68.9%) remained LoF/likely LoF after curation (Figure 2A, Table S1). The frequency of pLoF evasion and potential technical artifacts in AR genes was significantly lower than the 66.5% evasion rate observed for heterozygous pLoF variants in dominant disease-genes (n=403),^18^ where LoF variants are expected to be absent or depleted from gnomAD (p=5.44*10^−43^; Supplementary S10).

**Figure 2:**
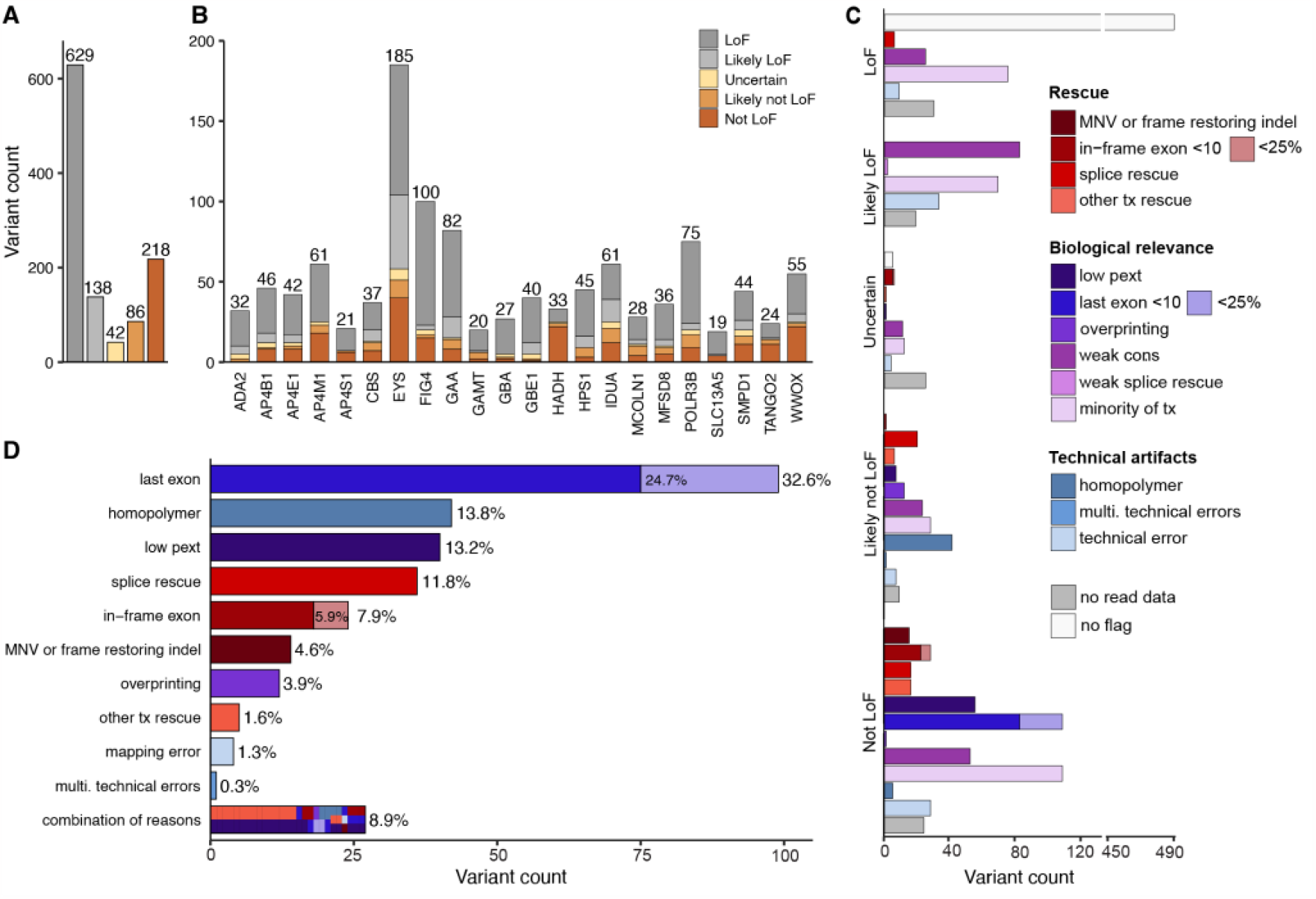
Evaluation of 1,113 heterozygous high-confident predicted loss of function (pLoF) variants in 22 recessive disease genes in gnomAD predict 27.3% do not result in true LoF because of rescue by secondary sequence properties, uncertain biological relevance, or potential technical artifacts. (A) Distribution of LoF verdicts in whole set, (B) and per gene. (C) The number of pLoF variants assigned with each flag within each classification category, colored by categories: potential technical artifacts (blue), uncertain biological relevance (purple), and rescue by secondary sequence properties (red). (D) Reasons for variants predicted as likely not LoF/not LoF (n = 304 variants). Combination of reasons refers to variants with more than one reason for likely not LoF/not LoF verdict. Tx: transcript, multi: multiple.

A variable proportion of LoF evasion and potential technical artifacts was observed between genes, explained by gene-specific properties (Figure 2B). *HADH* displayed the highest degree of LoF evasion and potential technical artifacts, 72.7% (24 of 33 variants), mainly due to several regions with low pext scores, while *GBE1* displayed the lowest degree (5.0%, 2/40 variants). Across the 1,113 variants, the most common variant class was frameshift, with 470 variants (42.2%), followed by 360 stop-gained/nonsense variants (32.3%), and 283 (25.4%) essential splice variants (153 donors (13.8%) and 130 acceptor variants (11.7%) (Supplementary Fig. 11A-C). The proportion of variants that were predicted to not result in LoF depended on variant class (Supplementary Figure S11D). Stop-gained/nonsense variants were most likely to be predicted as true LoF compared to other variant types (p=4.01*10^−6^; post hoc 2×2 chi-squared test; Bonferroni significance threshold p<.0125 for 4 post hoc tests) with only 19.1% predicted as likely not LoF/not LoF, whereas 40.0% of essential splice acceptor variants were predicted as likely not LoF/not LoF (p=2.81*10^−4^, post hoc 2×2 chi-squared test). Frameshift variants also had a slightly elevated proportion of predicted likely not LoF/not LoF (30.9%, p=0.0102, post hoc 2×2 chi-squared test). Essential splice donor variants did not significantly deviate from the mean across other variant types (24.8%, p=0.463, post hoc 2×2 chi-squared test).

Variants curated as likely not LoF/not LoF had a median of two flags per variant, (Supplementary Figure S12), though for the vast majority (91.1%, 277/304 variants) a single flag was sufficient to label them as likely not LoF/not LoF: last exon (32.6%), homopolymer (13.8%), low pext (13.2%), splice rescue (11.8%), in-frame exon (7.9%), MNV or frame-restoring indels (4.6%), overprinting (3.9%), other transcript rescues (e.g. translational reinitiation, overhang exon) (1.6%), mapping errors (1.3%), and multiple technical artifacts (0.3%) (Figure 2D). Variants interpreted as likely not LoF/not LoF due to their location, either in the last exon or at the border of an in-frame exon, were only considered likely not LoF/not LoF if they removed less than 25% of the coding sequence. Of 99 variants interpreted as likely not LoF/not LoF due to location in the last exon, 75 of 99 (75.7%) terminated in the last 10% of the coding sequence and the remaining 24 variants terminated in the last 10-25% of the coding sequence. A similar pattern was observed for splice variants at the border of in-frame exons; 18 of 24 variants (75.0%) resulted in a deletion of less than 10% of the protein coding sequence, and 6 variants resulted in a deletion spanning 10-25% of the protein coding sequence. Although the vast majority of variants had a single primary explanation for evasion (91.1%), they often had additional less impactful flags (Supplementary Figure S13). Only 27 variants (8.9%) were assigned multiple flags that are sufficient for a prediction of likely not LoF/not LoF, with the most prevalent combination being other transcript rescues (overhang exon) and low pext.

An analysis on the effect of all splice variants (n = 283) using SpliceAI revealed that 25.8% of splice variants are predicted to lead to a potential LoF rescue by an in-frame cryptic splice event, in-frame exon skipping, or location at a non-essential exon (Supplementary Figure S14). Upon further evaluation of intronic in-frame splice events, 6 variants were predicted to splice an intronic sequence that included a termination codon. These were expected to result in LoF, highlighting the need to consider several rescue mechanisms in parallel when assessing a final verdict. Other categories of splicing effects that retained LoF/likely LoF interpretations were out of frame rescues, out of frame exon skipping, multiple out of frame events, uncertain predictions, and intron retention.

Variants given a verdict of LoF/likely LoF had either no flags (white, Figure 2C), or had potential technical artifact and/or uncertain biological relevance flags (light blue and purple, Figure 2C) not considered strong enough evidence for the variant to be predicted as likely not LoF/not LoF (Table 1). The 42 variants (3.8%) that were given a verdict of uncertain LoF were mainly uncertain because of unavailable read data for visualization of potential frame-restoring indels in the surrounding region for variants in gnomAD. Of the 42 variants, 5 (11.9%) had no flags assigned to them, but were marked uncertain LoF mostly due to unclear splicing mechanisms.

### ACMG/AMP guided pLoF interpretation

The LoF curation protocol presented above predicts a variant’s likelihood to result in LoF but does not assess the variant’s pathogenicity. A pLoF variant curated as likely not LoF/not LoF may still be pathogenic via other mechanisms besides complete loss of gene expression, such as an in-frame deletion of a functional domain resulting in a catalytically inactive protein. Here we build upon the previous ClinGen SVI guidelines by Abou Tayoun et al.^4^ and provide a framework for further adjusting PVS1 for pLoF variants with a verdict of uncertain LoF, likely not LoF, or not LoF (Figure 3).

**Figure 3:**
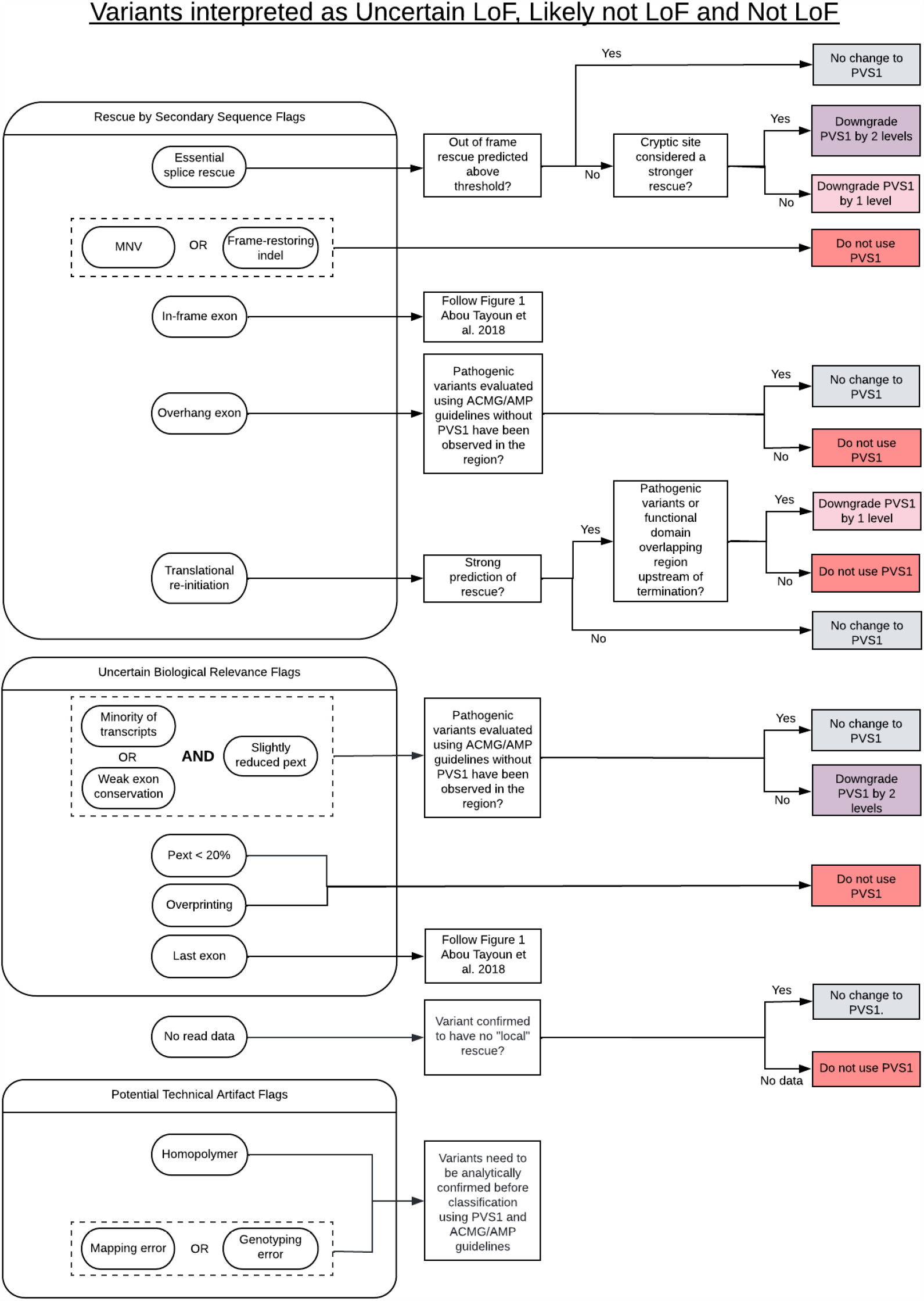
Framework for adjusting ACMG/AMP PVS1 criteria for variants curated as uncertain LoF, likely not LoF, or not LoF. Variants with technical artifact flags need analytical confirmation for ACMG/AMP classification using PVS1. For analytically confirmed uncertain LoF, likely not LoF, or not LoF variants, PVS1 should be modified accordingly; no changes to PVS1 (grey), downgrade PVS1 by one level (light pink, PVS1_strong max), downgrade PVS1 by two levels (purple, PVS1_moderate max), or not to use PVS1 at any level (dark pink). Downgrading is done by worst consequence and not in an additive manner.

The assessment of variants as technical artifacts is important for the accurate return of individual patients results, as well as the review of evidence from population databases such as gnomAD to ensure that variant occurrence and population allele frequencies are accurately represented. All variants assigned technical flags are by definition located in a region with quality concerns, and therefore allele frequencies in these regions in gnomAD may be higher than expected and thus, need to be interpreted with care.^12^ Variants with quality concerns should be analytically confirmed before assessing for pathogenicity, as only if the variant is real will it confer a disease risk. However, the analytic validity of a variant is a separate step from pathogenicity classification and therefore technical artifact consideration is not used to modify PVS1 strength in the context of a variant classification framework.

Figure 3 highlights modifications to the application of PVS1 following use of this framework for further pLoF interpretation. If a variant has been assigned several flags that suggest downgrading PVS1, the flag resulting in the most substantial downgrade should be applied to the curation (and not the sum of different consequences). For example, if a curation has resulted in both a splice rescue flag (downgrade PVS1) and a low pext flag (do not use PVS1), then PVS1 should not be applied, instead of downgrading.

Evaluation of the 479 variants (of the 1,113 assessed) that had ClinVar entries demonstrated that the 125/479 pLoF variants that were predicted as likely not LoF/not LoF were more likely to be classified as B/LB (16/125, 12.8%) in ClinVar compared to the 346/479 pLoF variants predicted as LoF/likely LoF (2/346, 0.6%) (p <0.0001, Fisher’s exact test). A full comparison of all LoF curations compared to ClinVar pathogenicity classifications can be seen in Supplementary Table S2. Of note, the accuracy of the ClinVar pathogenicity classifications was not formally evaluated.

To investigate the concordance between variants predicted to evade LoF, and the effect on PVS1, we assessed a subset of 200 out of 304 pLoF variants curated as likely not LoF/not LoF (Figure 3, Supplementary Table S3). Variants flagged as potential artifacts cannot be assessed using PVS1 unless analytically confirmed, therefore 37 variants with a technical artifact flag resulting in likely not LoF/not LoF verdict were filtered out, leaving 163 variants. Including the updates to PVS1 presented in this manuscript, the PVS1 criteria was downgraded by at least 1 level, to PVS1_strong or lower, for 162 of 163 variants (99.4%) curated as likely not LoF/not LoF. Of the 162 downgraded variants, PVS1 was affected in 17.2% (28 variants) due to updated guidelines of this framework with an effect on the final ACMG/AMP classification for 20 of those 28 variants (19 variants downgraded from likely pathogenic to VUS, and one variant from pathogenic to likely pathogenic).

### DISCUSSION

There are several mechanisms by which pLoF variants can escape LoF, which is why careful assessment beyond standard annotation pipelines are key to reduce false positive rates and ensure accurate prediction in both research and clinical settings. We present a new framework that refines the interpretation of pLoF variants’ predicted impact and introduce a structured methodology to predict variants as LoF, likely LoF, uncertain LoF, likely not LoF, or not LoF. Further, we expand on how this evidence ties into the assessment of pathogenicity by current ACMG/AMP guidelines, and specifically how PVS1 should be modified, in line with and further building upon standards provided by the ClinGen SVI working group.^4^

The LoF curation protocol introduces three different categories of evidence that should be assessed: rescue by secondary sequence properties, uncertain biological relevance, and potential technical artifacts. Of 1,113 high-confidence pLoF variants in 22 AR disease genes investigated here, 27.3% were predicted as likely not LoF/not LoF. The main reasons were truncation in the 3’ end of the gene, location in a homopolymer region, location in a low pext region, and essential splice variants with in-frame cryptic rescues, highlighting the importance of detailed assessment to accurately interpret a pLoF variant. A similar rate of LoF evasion and potential technical artifacts is observed for homozygous pLoF variants in gnomAD, in line with the expected enrichment of rescue mechanisms and artifacts seen for pLoF variants in general.^7,16^ As expected, this rate of pLoF evasion and potential technical artifacts is higher for heterozygous pLoF variants in haploinsufficient genes associated with severe disorders not expected in population databases like gnomAD (66.5%, Supplementary Figure S10).^18^

In addition to the expected difference in evasion between gene sets, there were differences in frequency of evasion between LoF variant classes. Stop-gained variants were more likely to be predicted as true LoF with a lower evasion rate of 19%, while 40% of essential splice acceptor variants were predicted likely not LoF/not LoF. The elevated evasion rate for essential splice acceptor variants was mainly driven by location in the last exon and cryptic splice site rescue, with the cryptic splice acceptor variant rescues suggestively being due to less consensus site conservation for splice acceptor than donor sites.^34,56^ This can guide how we think of splice acceptor variants as escape seems more common, whereas nonsense variants to a large extent are predicted as true LoF (Supplementary S11).

The LoF curation protocol does not determine variant pathogenicity, but rather predicts the likelihood of a pLoF variant evading LoF or acting as a potential technical artifact. Additional information regarding variant classification is required to determine a pLoF variant’s pathogenicity, including if there is a non-NMD or other less damaging predicted effect of a pLoF variant, as well as segregation data, case-level evidence, functional evidence, *de novo* evidence, and population evidence. Variants predicted as likely not LoF/not LoF were enriched for variants classified as benign/likely benign in ClinVar, highlighting that our framework can identify variants that potentially evade LoF and do not cause disease. The two variants predicted as likely LoF through our framework, but classified as benign/likely benign in ClinVar (indicating potential missed evasion of LoF by our framework) are annotated as pLoF in one non-MANE Select IDUA transcript (ENST00000247933.4) (three transcripts reported in Gencode v19), with a mean pext score of 0.4 (max for the gene is 0.6 - 0.7), which suggests some biological relevance and the variants were therefore not excluded as likely not LoF/not LoF across all tissues. One can speculate that the affected transcript is non-essential for the enzymatic function of *IDUA* that is disrupted in the IDUA-associated metabolic condition Mucopolysaccharidosis (OMIM #607014).

Importantly, variants predicted as likely not LoF/not LoF by our framework may still be pathogenic via mechanisms other than loss of protein expression. In particular, the 89 variants predicted as likely not LoF/not LoF that were classified as pathogenic/likely pathogenic warrant further scrutiny to confirm their pathogenicity is due to other evidence besides a loss of function mechanism. We established an effect on PVS1 in 99.4% (162 of 163) of assessed variants predicted as likely not LoF/not LoF using the existing PVS1 guidelines^4,11^ in combination with our framework. Importantly, PVS1 was affected in 17.2% (28 of 163) of variants as a result of the updated guidelines provided in this report, mostly due to essential splice site variants predicted to be rescued by in-frame cryptic splice events, MNVs, or frame-restoring indels. This result highlights the importance of considering the new properties presented here when assessing pLoF variants and their pathogenicity. Further, for 71.4% (20 of 28) variants, the effect on PVS1 resulted in a downgraded ACMG/AMP variant classification, mostly from likely pathogenic to uncertain significance, highlighting the clinical impact of this framework. Not considering mechanisms of escape confers a risk of overestimating the pathogenicity of pLoF variants.

One important consideration during LoF curation is that the general rate of evasion and potential technical artifacts (27.3% in this variant set) will vary depending on how the variants were ascertained (from patient or in population data), as well as methods used for identifying the variants (large-scale sequencing or standardized clinical sequencing including orthogonal confirmation). Since LoF variants as a group are under negative selection and the proportion of artifacts from genotyping will be constant across the genome and variant classes, pLoF variants in population data will be more enriched for artifacts, especially in disease genes constrained for LoF.^13,16^ In patient cohorts enriched with individuals affected by severe disease, the contrary is true, with an expected enrichment for pLoF variants that are true positives and also pathogenic. Therefore, it is expected that the evasion rate and number of technical artifacts are much lower in a patient cohort. Thus, the source of the variant data should impact the conservative or lenient threshold set for the different flags presented in this protocol, with a more conservative approach being recommended for any curation of population data.

This protocol aims to include any of the community established and accepted mechanisms of LoF evasion and predictions of technical artifacts to improve pLoF predictions. However, additional mechanisms resulting in LoF evasion have been suggested and are likely to be established in the future. For example, it has been suggested that pLoF variants in an exon longer than 400 bp, or a variant located in the first 150 coding bp will escape nonsense mediated decay.^6^ Genes susceptible to clonal hematopoiesis due to proliferative advantage from haploinsufficiency (monoallelic LoF) is an aspect not within the scope of this protocol (beyond hard filtering variants with an allele balance of less than 20%). However, clonal hematopoiesis should be considered when assessing pLoF variants in population databases in genes associated with this phenomenon.^57,58^

In conclusion, we present a framework that aids in the interpretation of pLoF variants by considering mechanisms of LoF evasion and indications of potential technical artifacts, alongside updated guidelines for applying PVS1 for classifying pLoF variant pathogenicity. The results presented here highlight how inadequate pLoF variant assessment stands a risk of overinterpreting the effect and pathogenicity of pLoF variants and that this framework can substantially reduce the false positive rate of pLoF in both research and clinic settings.

## Data Availability

All data produced in the present work are contained in the manuscript

## ACKNOWLEDGEMENTS

We thank the individuals and researchers whose data is in gnomAD for their contributions to research. We thank the ClinGen Sequence Variant Interpretation Working Group for useful discussion of PVS1 modifications and Raymond Walters for valuable advice on statistical analysis. This work was supported in part by the following NIH grants: NHGRI U24HG011450, NHGRI U24HG006834, NHGRI U01HG011755, and NHLBI R01HL143295, and by research funding from the Chan Zuckerberg Initiative to H.L.R. and from Genzyme Corporation to D.G.M. S.G. was supported by The Knut and Alice Wallenberg Foundation scholarship program for postdoctoral studies at the Broad Institute. Its contents are solely the responsibility of the authors and do not necessarily represent the official views of NIH, CZI, or Genzyme.

## DECLARATION OF INTERESTS

S.H. is an employee of Ambry Genetics. D.G.M. is a founder with equity of Goldfinch Bio, serves as a paid advisor to GSK, Variant Bio, Insitro, and Foresite Labs, and received research funding from Genzyme. H.L.R. receives funding from Illumina to support rare disease gene discovery and diagnosis. A.O-D.L. is a paid advisor of Tome Biosciences, Congenica, and the Simons Foundation SPARK for Autism study.

