## Supplemental Files for "Advanced variant classification framework reduces the false positive rate of predicted loss of function (pLoF) variants in population sequencing data"

### SUPPLEMENTARY MATERIAL

**Advanced classification framework demonstrates a large fraction of population predicted loss of function (pLoF) variants in genomic sequencing do not result in LoF**

Moriel Singer-Berk[1,2,*], Sanna Gudmundsson[1,2,3,4*], Samantha Baxter[1,2], Eleanor G. Seaby[1,2,3,5], Eleina England[1,2,3], Jordan C. Wood[1,2], Rachel G. Son[1], Nicholas A. Watts[1], Konrad J. Karczewski[1,2], Steven M. Harrison[1,6], Daniel G. MacArthur[1,7,8], Heidi L. Rehm[1,2], Anne O'Donnell-Luria[1,2,3]

1. Program in Medical and Population Genetics, Broad Institute of MIT and Harvard,

Cambridge, MA, USA

2. Center for Genomic Medicine, Analytic and Translational Genetics Unit, Massachusetts General Hospital, Boston, MA, USA

3. Division of Genetics and Genomics, Boston Children's Hospital, Harvard Medical School, Boston, MA, USA

4. Science for Life Laboratory, Department of Gene Technology, KTH Royal Institute of Technology, Stockholm, Sweden.

5. Genomic Informatics Group, University Hospital Southampton, Southampton, United Kingdom

6. Ambry Genetics, Aliso Viejo, CA, USA

7. Centre for Population Genomics, Garvan Institute of Medical Research and UNSW Sydney, Sydney, New South Wales, Australia

8. Centre for Population Genomics, Murdoch Children's Research Institute, Melbourne, Australia

*equal contribution

#### SUPPLEMENTARY TABLES

**Table S1:** 1,113 variants manually classified using this advanced pLoF framework, including final verdict and error flags applied.

**Table S2:** List of variants predicted as likely not LoF/not LoF that have an effect on PVS1 (downgrade vs upgrade) and variant pathogenicity classification.

**Table S3:** Number of variants with at least one ClinVar submission grouped by predicted LoF effect following our protocol and ClinVar pathogenicity classification.

| | Benign/ Likely benign | VUS/ Conflicting | LP/ P | Not Provided | Total |
|---|---|---|---|---|---|
| LoF/likely LoF | 2 | 19 | 324 | 1 | 346 |
| Uncertain LoF | 0 | 1 | 7 | 0 | 8 |
| Likely not LoF/not LoF | 16 | 20 | 89 | 0 | 125 |
| Total | 18 | 40 | 420 | 1 | 479 |

#### SUPPLEMENTARY FIGURES

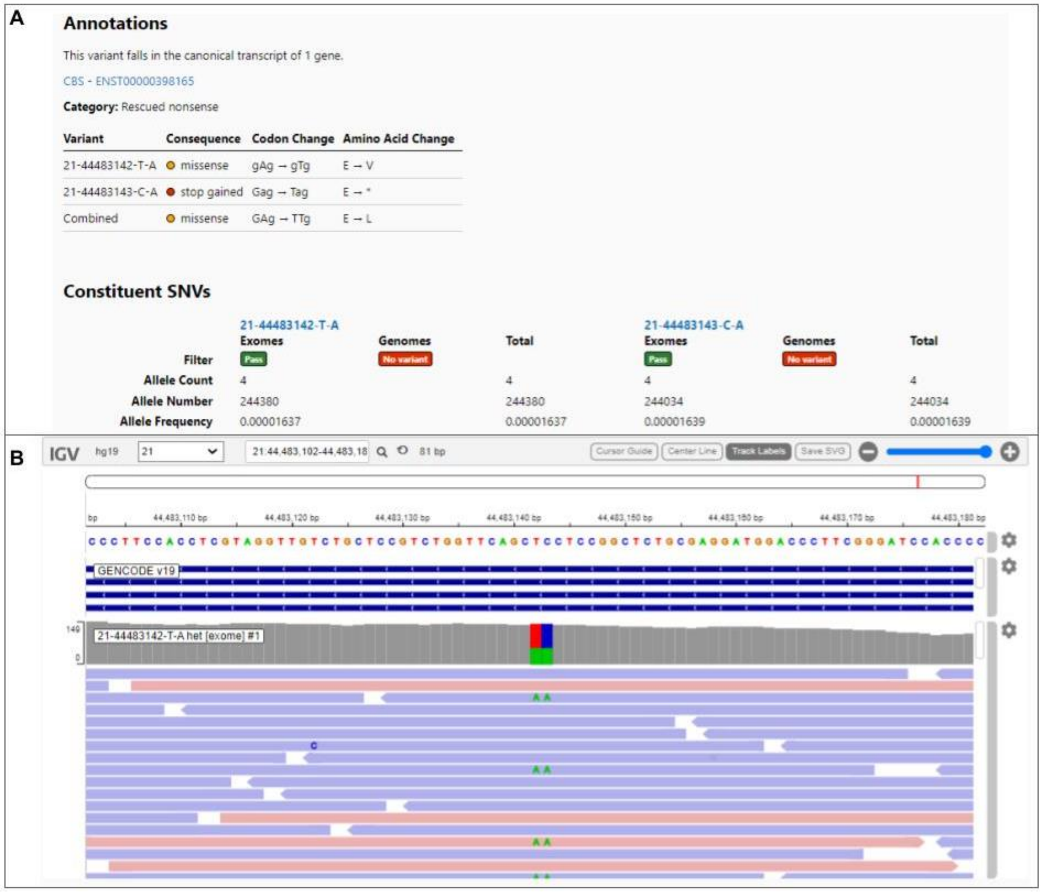


**Supplementary Figure S1:** Multi-nucleotide variant. (A) Two variants annotated individually in gnomAD as missense and stop gained respectively. The combined effect can be noted to result in a different missense variant. (B) Read data from IGV in gnomAD that displays both SNVs are in-phase.

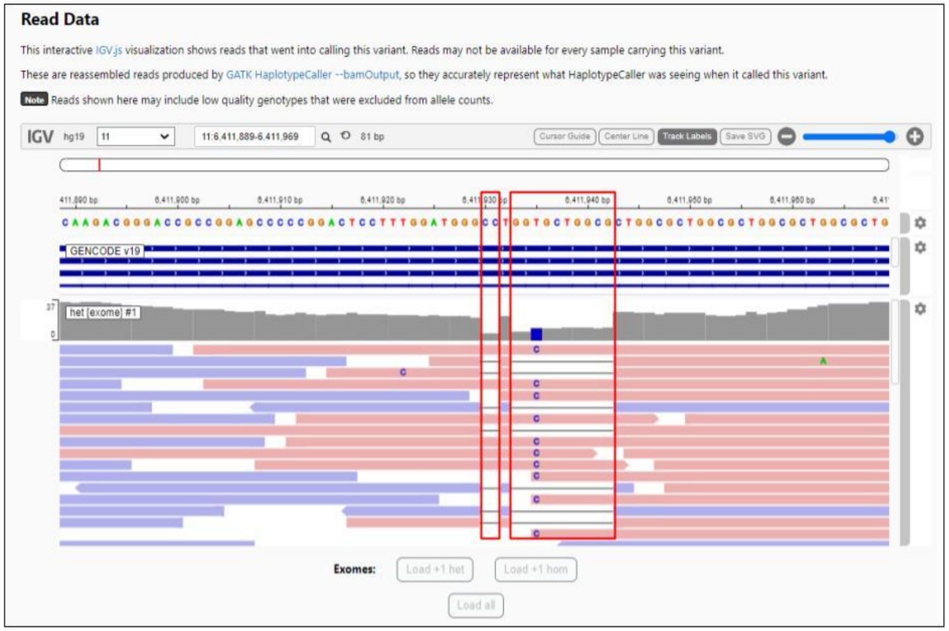


**Supplementary Figure S2:** Frame-restoring indel. The two individual variants (both highlighted in red boxes) are in cis deletions of 2 bp and 10 bp, respectively. The sum of these deletions (12 bp, 4 amino acids) is in-frame. Typically, this is a single mutational event that is inaccurately represented by the mapping and calling algorithms as two (or more) events.

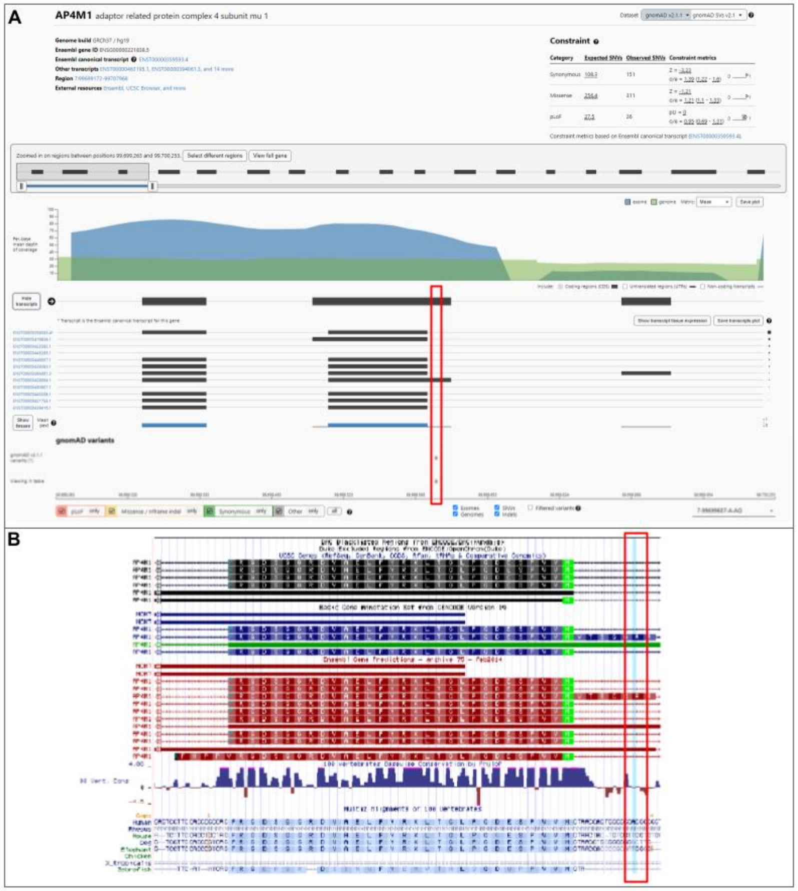


**Supplementary Figure S3:** Variant with overhang exon flag in *AP4M1*. Variant position highlighted (red box). (A) The zoomed in region of *AP4M1* exons 1-3 shows regions of exon 2 where there is an overhang as well as low pext scores relative to the maximum for the gene. (B) UCSC genome browser clearly shows the region containing the pLoF variant is in an overhanging part of a single transcript, and the overhanging region is weakly conserved in vertebrates.

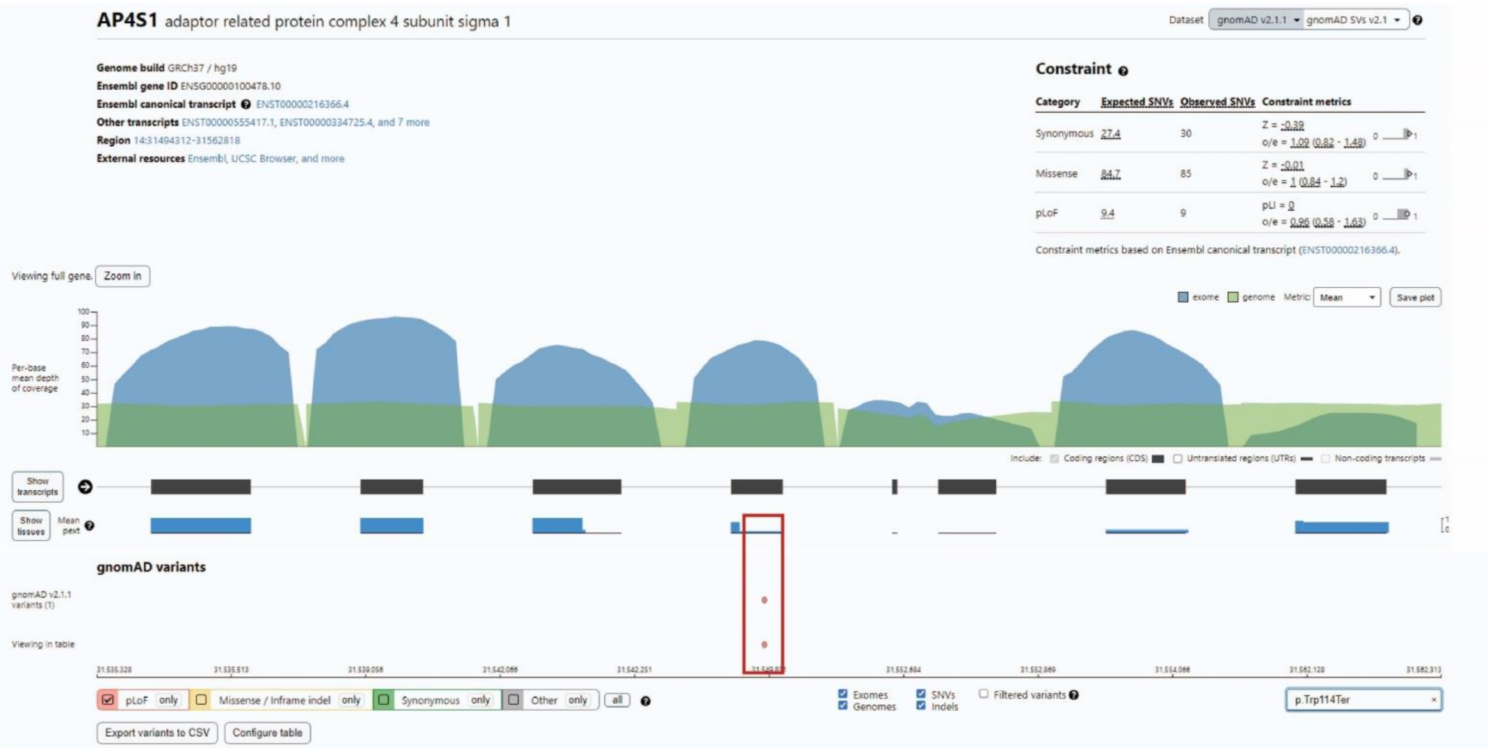


**Supplementary Figure S4:** Example of a variant falling in a low pext region (red box). The variant is identified in the gnomAD variant track as a pLoF variant falling in exon 4 of 6 for the canonical transcript. The pext score at this position of the exon (0.1) is much lower than the maximum score for the gene (1).

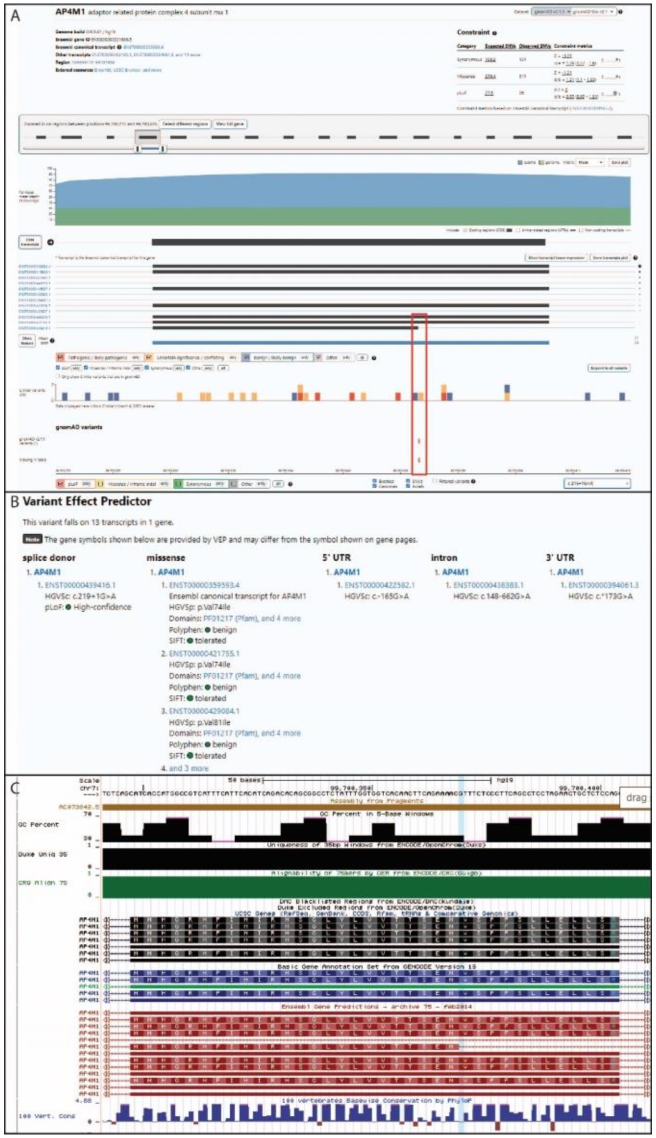
A
AP4M1 adaptor related protein complex 4 subunit mu 1
Constraint
gnomAD variants
B Variant Effect Predictor
This variant falls on 13 transcripts in 1 gene.
Note The gene symbols shown below are provided by VEP and may differ from the symbol shown on gene pages.
splice donor
1. AP4M1
1. ENST00000439416.1
HGVSc: c.219+1G>A
pLoF: High-confidence
missense
1. AP4M1
1. ENST00000359593.4
Ensembl canonical transcript for AP4M1
HGVSp: p.Val74Ile
Domains: PF01217 (Pfam), and 4 more
Polyphen: benign
SIFT: tolerated
2. ENST00000421755.1
HGVSp: p.Val74Ile
Domains: PF01217 (Pfam), and 4 more
Polyphen: benign
SIFT: tolerated
3. ENST00000429084.1
HGVSp: p.Val81Ile
Domains: PF01217 (Pfam), and 4 more
Polyphen: benign
SIFT: tolerated
4. and 3 more
5' UTR
1. AP4M1
1. ENST00000422582.1
HGVSc: c.-165G>A
intron
1. AP4M1
1. ENST00000438383.1
HGVSc: c.148-662G>A
3' UTR
1. AP4M1
1. ENST00000394061.3
HGVSc: c.*173G>A
C
Scale
chr7:
50 bases
hg19
99,700,350
99,700,400
drag
GC Percent in 5-Base Windows
GC Percent
Duke Uniq 35
100 Vert. Cons

**Supplementary Figure S5:** Example of a pLoF variant annotated as a splice donor variant not supported by per base expression (pext) score. (A) *AP4M1* gnomAD gene page with variant location across Ensembl transcripts (red box). A consistent pext score across the splice site (no drop at the splice site) suggests splicing in a transcript of low biological relevance (low/no expression). (B) The variant is annotated as a missense variant in all but one transcript where it is annotated as pLoF (C) UCSC genome browser confirms that the entire exon is conserved.

**Commented [1]:** I cant see B, and also A is very poor quality. the details referred to in the figure text of B is not readable?

**Commented [2]:** I'd split B and C to two independent rows to be able to make them bigger (but also need to be higher quality, prep the figures in illustrator at a with of A4 and 300dpi)

**Commented [3]:** Agree with this - if you want to keep C, then I'd make B and C stacked/full width of the figure to make it easier to see

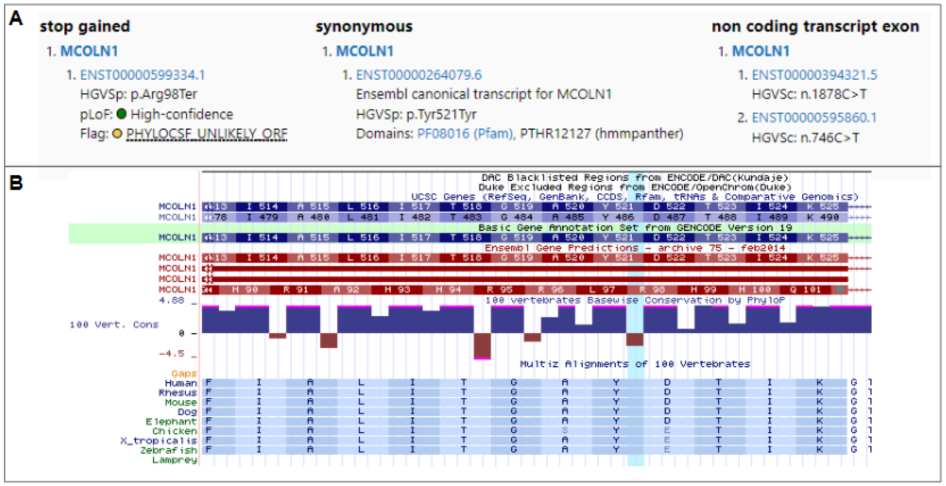


**Supplementary Figure S6:** Example of a pLoF variant with overprinting flag. (A) This variant in *MCOLN1* is annotated as p.Arg98Ter (stop gained) in one transcript and as p.Tyr521Tyr (synonymous) in the canonical transcript. (B) The UCSC browser displays weak conservation of the wobble position in the canonical transcript, where the variant is annotated as synonymous, but not for the alternative transcript, hence, the variant is not predicted pLoF in the most biologically relevant transcript.

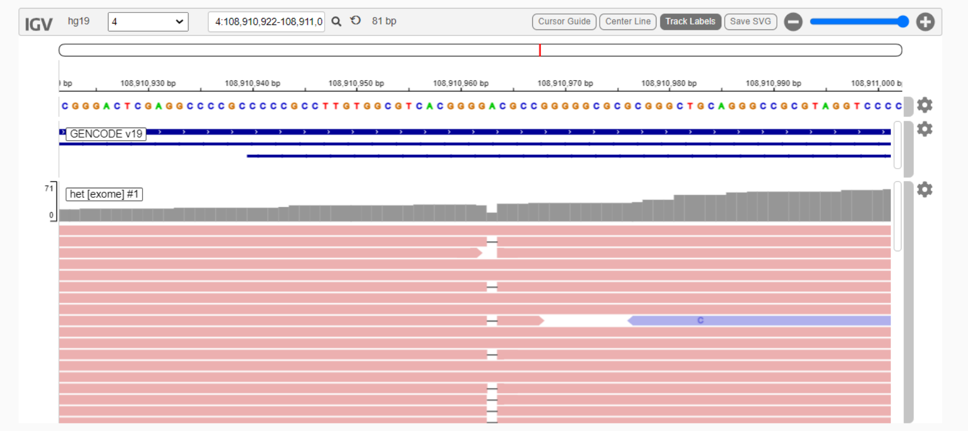


**Supplementary Figure S7:** Example of strand bias in IGV read data. Deletion of a single nucleotide base is called only in reverse strands from sequencing (pink).

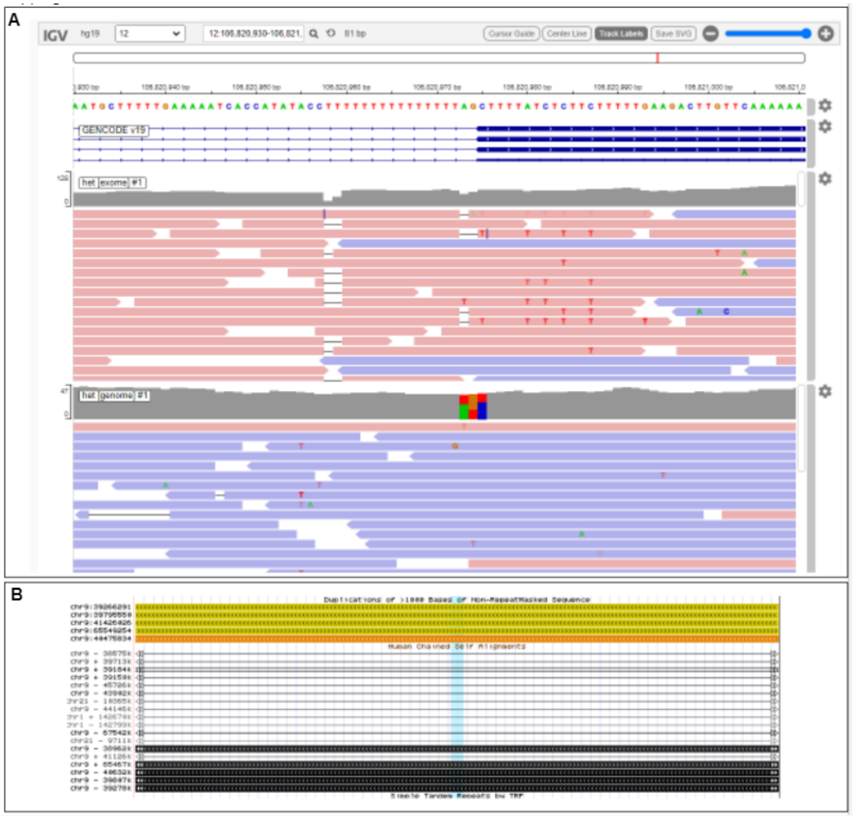

**Supplementary Figure S8:** Example of mapping flag from LoF curation protocol. (A) Evidence of complex mapping from sequencing data resulting in multiple variant calls at varying allele balance across a small region of the genome. (B) Additional evidence towards mapping difficulties in sequencing as seen in UCSC genome browser mapping tracks “Segmental Duplications” and “Self Chain” (Repeats section).

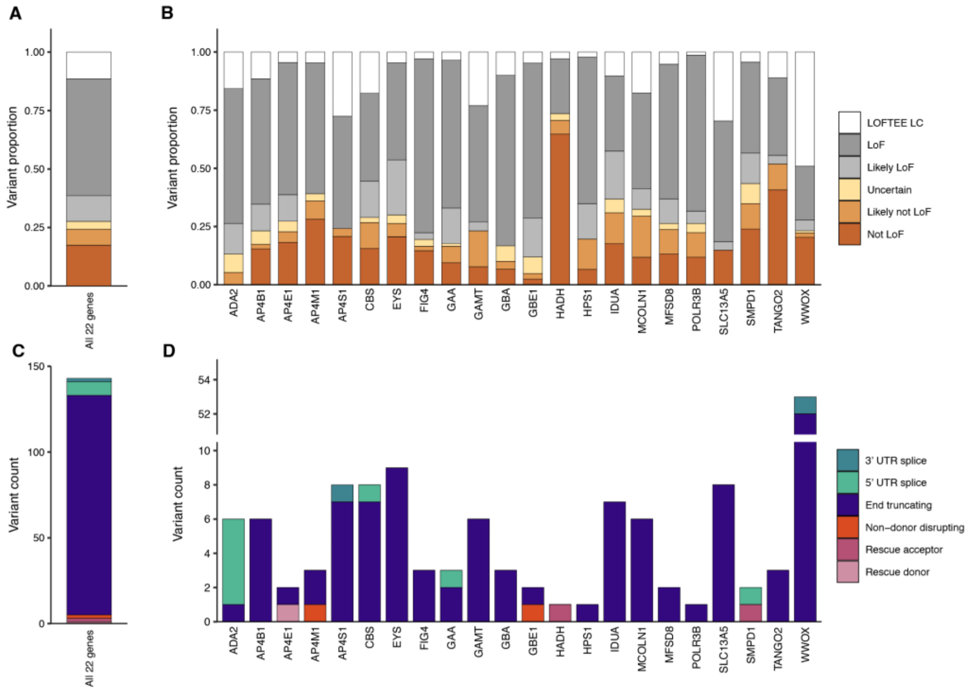


**Supplementary Figure S9:** Proportion of LoF predictions. (A-B) Frequency of coding pLoF variants (n=1256) filtered by each method, and across all 22 recessive disease genes combined (A), and per gene (B). (C-D) Variants filtered by the Loss-Of-Function Transcript Effect Estimator (LOFTEE; n=143), and across all 22 recessive disease genes combined (C), and per gene (D).

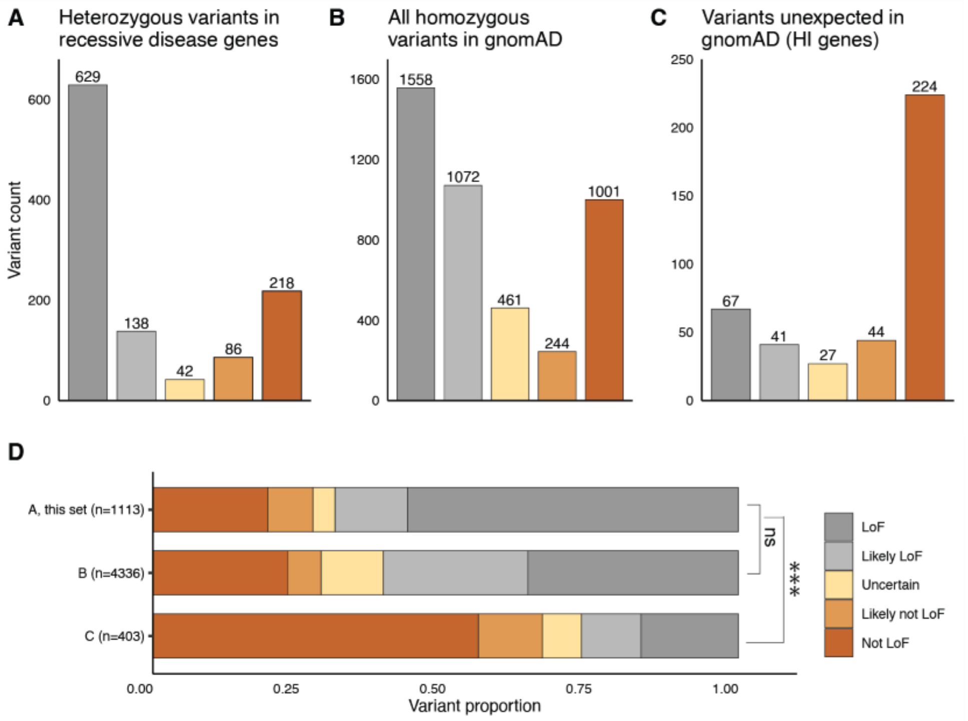


**Supplementary Figure S10:** Predicted loss of function (pLoF) curation outcome in different genesets, revealed similar proportion of variants predicted as likely not LoF/not LoF between this set (heterozygous pLoF in autosomal recessive (AR) disease genes) and all homozygous pLoF variants in gnomAD, 27.3% vs 28.7% (p=0.37, Fisher's exact test). As expected pLoF variants in haploinsufficiency (HI) genes associated with autosomal dominant (AD) disease (not expected, or expected to be depleted from gnomAD) display a higher rate of likely not LoF/not LoF predictions of 66.5% (32.2% vs. 66.5%, $p=5.44*10^{-43}$, Fisher's exact test). ***: p<0.0001. Ns: non-significant.

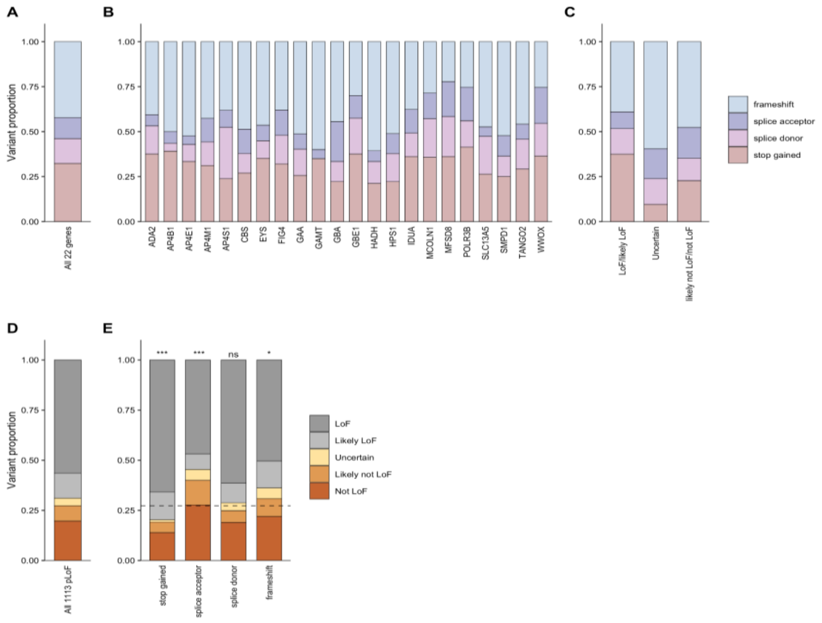


**Supplementary Figure S11:** Proportion of each pLoF variant type in all 22 recessive disease genes combined (A), per gene (B), and per verdict group (C). Of 1,113 variants, 27.3% were predicted likely not LoF/not LoF (D). The proportion of variants that were predicted to not result in LoF depended on variant type. Stop-gained variants were most likely to be predicted as true LoF compared to other variant types ($p=4.01*10^{-6}$; post hoc 2x2 chi-squared test; Bonferroni significance threshold p<.0125 for 4 post hoc tests) with only 19.1% predicted as likely not LoF/not LoF, whereas 40.0% of splice acceptor variants were predicted as likely not LoF/not LoF ($p=2.81*10^{-4}$, post hoc 2x2 chi-squared test). Frameshift variants also had a slightly elevated proportion of predicted likely not LoF/not LoF (30.9%, p=0.0102, post hoc 2x2 chi-squared test). Splice donor variants did not significantly deviate from the mean (27.3%, dash line) across other variant types (24.8%, p=0.463, post hoc 2x2 chi-squared test). (E).

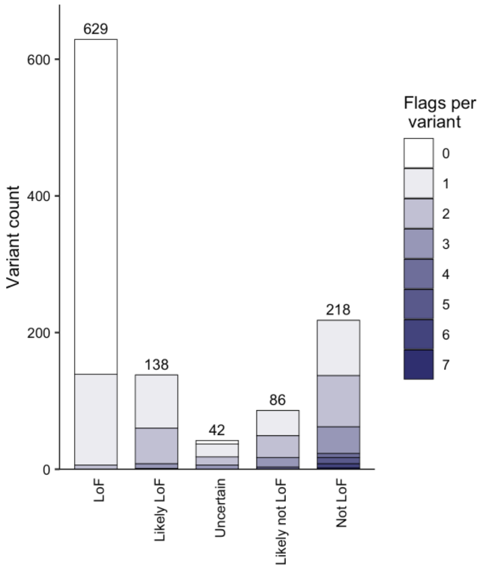


**Supplementary Figure S12:** Number of flags per variant per verdict category.

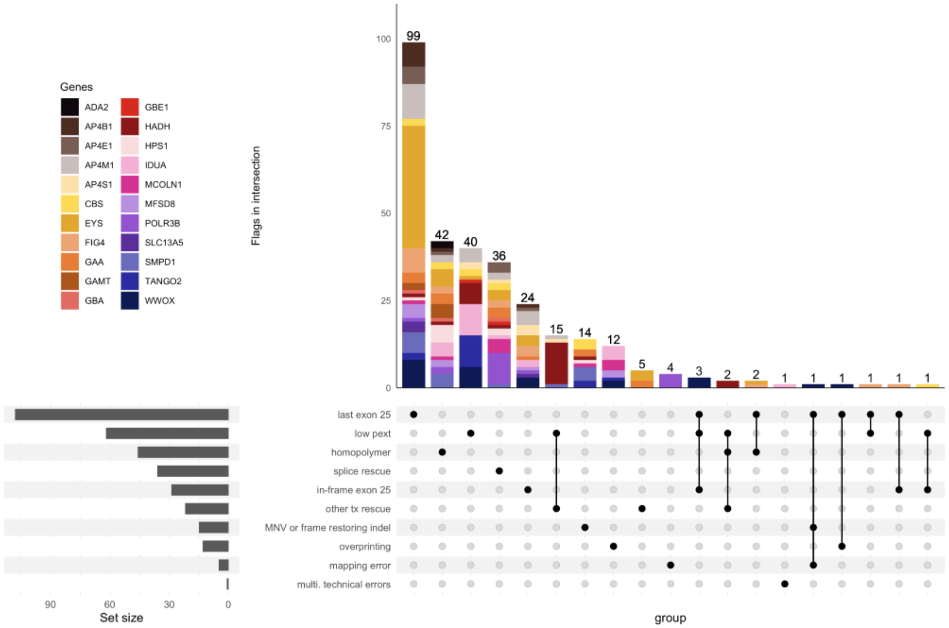


**Supplementary Figure S13:** Reasons for 304 of 1,113 variants predicted as likely not LoF/not LoF, colored by gene the variant occurred in.

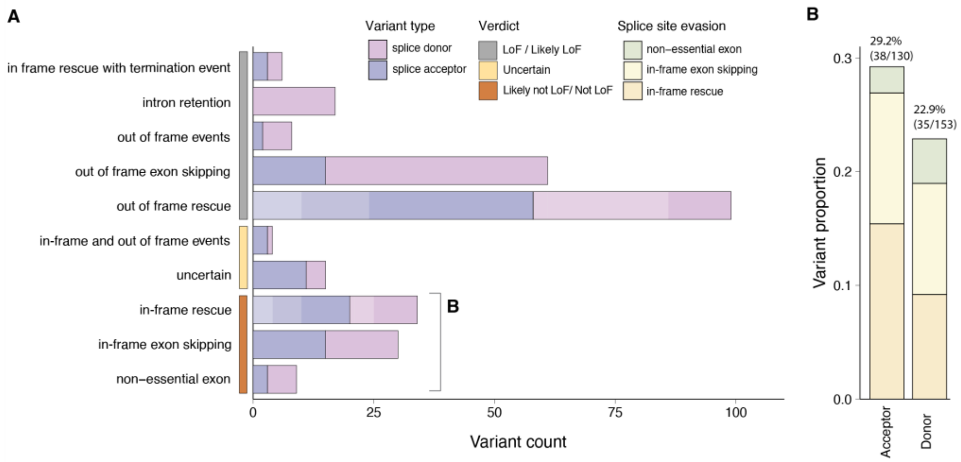


**Supplementary Figure S14:** SpliceAI predictions. Predicted effect of all splice site variants (n = 283) using SpliceAI (A). 29.2% (38/130) of acceptor and 22.9% (35/153) of donor variants are predicted as likely not LoF/not LoF (B).
